# Parent and physiotherapist perceptions about movement skills of young children with juvenile idiopathic arthritis

**DOI:** 10.64898/2026.06.10.26355384

**Authors:** Elyse Letts, Julie Herrington, Michelle Batthish, Chloe Bedard, Emily Bremer, Jan Willem Gorter, Sara King-Dowling, Joyce Obeid

**Affiliations:** Child Health & Exercise Medicine Program, Department of Pediatrics, McMaster University, 1280 Main St W, Hamilton, ON, L8S 4L8, Canada; School of Rehabilitation Science, McMaster University, 1280 Main St W, Hamilton, ON, L8S 4L8, Canada; McMaster Children’s Hospital, 1280 Main St W, Hamilton, ON, L8S 4L8, Canada; Department of Pediatrics, Division of Rheumatology, McMaster University, 1280 Main St W, Hamilton, ON, L8S 4L8, Canada; Grand Erie District School Board, Brantford, ON N3T 5V3, Canada; School of Kinesiology, Acadia University, Wolfville, Nova Scotia, B4P 2R6, Canada; CanChild, Department of Pediatrics, McMaster University, 1280 Main St W, Hamilton, ON, L8S 4L8, Canada; Department of Pediatrics, The Children’s Hospital of Philadelphia, 3500 Civic Center Blvd, Philadelphia, PA 19104, United States of America; Department of Kinesiology, McMaster University, 1280 Main St W, Hamilton, ON, L8S 4L8, Canada

**Keywords:** JIA, qualitative, fundamental motor skills

## Abstract

**Objective:** The onset of juvenile idiopathic arthritis (JIA) in the early years (≤5 years) may negatively impact movement skill (encompassing related concepts of gross motor skills, fundamental movement skills, and functional ability) development. Few studies have explored the perceptions and needs of parents and physiotherapists towards children’s difficulty with these movement skills, essential to identify potential areas for added support. The objective of this study is to understand the perceptions of physiotherapists and parents towards movement skills of children with JIA.

**Methods:** Seventeen parents and 24 physiotherapists completed an online questionnaire consisting of multiple choice and open-ended questions about the movement skills of young children with JIA. Demographic and multiple choice questions were quantitively analysed using descriptive statistics. Open-ended responses were analyzed using qualitative conventional content analysis.

**Results:** About half (47%) of parents perceived their children to have movement difficulties, and 75% of physiotherapists described the movement skills of children with JIA as worse than other children of the same age. Our qualitative analysis revealed three general themes including: functional task difficulties; clinical variability in movement skills; and psychosocial components of movement skill difficulties.

**Conclusion:** This study provides an analysis of perceptions of physiotherapists and parents towards the movement skills of young children with JIA. A significant proportion of parents and physiotherapists identify movement difficulties among children with JIA that impact daily life. Future interventions co-designed with both parents and care providers targeting movement skills are needed.

**Significance and Innovation:**

- This study provides an analysis of perceptions of physiotherapists and parents towards the movement skills of young children with JIA
- About half (47%) of parents perceived their children to have movement difficulties, and 75% of physiotherapists described the movement skills of children with JIA as worse than other children of the same age
- Our qualitative analysis revealed that young children with JIA have functional task difficulties; that they experience clinical variability in movement skills; and that psychosocial components of movement skill difficulties impact both parents and children (e.g., concern around risk of flares with activity, frustration in not being able to participate)

## Introduction

Juvenile idiopathic arthritis (JIA) is the most common childhood rheumatic disease and is associated with joint swelling, pain, and stiffness.^1,2^ It impacts approximately 1 in 1000 Canadian children^3^ and is commonly diagnosed between ages 1-9 years.^4^ When symptom onset begins in the early years (under 5 years of age), it may have a substantial impact on movement skills.^5^ The early years are a critical time for the development of movement skills, a global term^6^ which encompasses related concepts of gross motor skills, fundamental movement skills, and functional ability (ability to perform activities that meet one’s basic needs and maintain health and wellbeing^7^). Some examples of movement skills include crawling, standing, balancing, walking, running, jumping, catching, throwing, and drawing.^8,9^ In children with JIA, the presence of joint pain and stiffness may lead to avoidance of activities that use the affected limb(s), and/or modifying movement patterns to compensate, which may reduce opportunities for developing these movement skills.^10^

A recent scoping review^11^ examined movement skills of children with JIA and found only eight studies that used a direct observation motor assessment and only five of these included children in the early years. Notably, all five studies^5,10,12–14^ found worse movement skills in children with JIA compared with age-matched norms or healthy peers. Given the ever-growing volume of research investigating health and wellbeing in JIA, it is somewhat surprising that so few studies have examined movement skill development in this population. With this limited knowledge base, more research is needed to better to understand which aspects of motor skill development are most impacted and of greatest concern for children, parents, and clinicians.
Parents of young children are often the first to notice symptoms (e.g., pain, swelling, avoidance) when they appear in the early years, prompting them to seek support.^15^ In the course of a diagnosis of JIA and subsequent treatment, families may also be referred to physiotherapy^16,17^ to help with muscle strength, joint mobility, core stability, and movement skills. Understanding how physiotherapists perceive movement skills among patients (e.g., what factors are of the most concern) can help us understand which elements of movement skills may be most impacted in JIA, highlighting areas for intervention, and in turn, help inform better clinical practice. To date, no studies have investigated the perceptions of physiotherapists on the movement skills of children with JIA.

Similarly, it is unclear if parents recognize any deficits or limitations in movement skills in their child with JIA. Parent perceptions of children’s participation in physical activity,^14,18^ experiences in caring for young children with JIA,^15^ and perceptions towards specific treatments (e.g., medication, physiotherapy)^19–21^ in children with JIA have been explored, however, to our knowledge, no study has elicited their perspectives of movement skills. Understanding parent perceptions of movement skills can help better guide clinical priorities and treatment as well as supporting the care needs of parents. For example, if many parents are unaware that their children may be behind on movement skill development, it may indicate the need for parent educational interventions about the importance of movement skills followed by a targeted movement skill intervention.
To address the limited evidence in the current literature, the objective of this study is to understand the perceptions of physiotherapists and parents towards the movement skills of children with JIA in the early years.

## Methods

We recruited parents and physiotherapists to complete an online questionnaire administered in REDCap.^22^ To be included, parents had to have a child aged 1-5 years with a diagnosis of JIA. Parents were recruited through the McMaster Children’s Hospital outpatient rheumatology clinic as well as through online advertising through a JIA organization (Cassie + Friends^23^). For physiotherapists, only those who had worked with children with JIA aged 1-5 years for at least 1 year were invited to participate. Physiotherapists were recruited through Canadian physiotherapy groups (e.g., Manitoba Physiotherapy Association), through a Canadian pediatric rheumatology allied health community of practice email list, and through word-of-mouth. Recruitment lasted from October 2024 through to October 2025. This study received ethics approval from the Hamilton integrated Research Ethics Board (HiREB# 9510) and all participants provided informed written consent.

### Questionnaire

Two versions of a questionnaire (one for parents and one for physiotherapists) were developed by the authors who have combined expertise in pediatric rheumatology, physiotherapy, motor development, and pediatric clinical research. The first section of the questionnaire collected relevant background and demographic information. For parents, this included relationship to the child, the child’s sex at birth, age, age at diagnosis, as well as affected joints. For physiotherapists, this included how many years they have worked with children with JIA as well as how many children with JIA they work with on average each month.

The remainder of the questionnaire questions covered four main topics: (i) General movement skills; (ii) upper body movement skills; (iii) lower body movement skills; and (iv) impact on daily living. Each section contained multiple choice (only one response allowed) questions (e.g., yes/no, none/a few/some/most/almost all) in addition to open-ended text responses. Open-ended text questions were provided in each section for both parents and physiotherapists (e.g., “Please describe and provide specific examples”). There was also one overarching open-ended text question at the end of the questionnaire for both parents and physiotherapists (e.g., “Is there anything else you’d like to share with us about the movement skills of your young patients/child with JIA?”). The full questionnaires for both parents and physiotherapists are available in supplementary material 1.

### Ages and stages questionnaire – 3^rd^ edition (ASQ-3)

For descriptive purposes, parents were asked to complete the gross motor subsection of the ASQ-3, also in REDCap, about their child. The ASQ-3 is a well-validated^24,25^ parent reported tool with 21 age-specific versions (from 1 through 66 months of age) and has been used in children with JIA.^26^ The gross motor subsection asks parents six questions for which parents rate their child as “yes” (performing the skill), “sometimes” (starting to perform the skill), or “no” (not yet performing the skill). Scores (possible range 0-60) are then compared to established age-specific thresholds and classified as typically developing (greater than 1-SD below the mean), the monitor zone (between 1- and 2-SD below the mean), and the warning zone (greater than 2-SD below the mean).

### Data analysis

Demographics, background information, and multiple choice questions were analysed using descriptive statistics. Open-ended text responses were analyzed using qualitative content analysis. Specifically, we used conventional content analysis^27^ with data managed in Dedoose.^28^ All open-ended text responses (total of 11 questions: 6 from parents, 5 from physiotherapists; details in supplementary material 1) were compiled, and one researcher (EL) coded the responses. The researcher first read through all responses and generated initial thoughts that were discussed with another research team member (JO) to generate an initial coding framework. EL then coded all excerpts in Dedoose and worked with JO to generate initial themes.^29^ These initial themes were then shared with the entire research team for discussion and refinement.

## Results

In total, 17 parents and 24 physiotherapists completed the questionnaire. Parents completing the questionnaire mostly identified as mothers (88%; n=15/17). Children had a mean ± SD (range) age of 3.4 ± 1.4 (1.4-5.3) years and were 88% female (n=15/17). They had a mean age at diagnosis of 2.3 ± 0.8 (1.2-3.7) years. Parents identified lower limb involvement in 16 (94%) children, upper limb involvement in 8 (47%) children, and spine involvement in 2 (12%) children. From the ASQ-3, the mean ± SD (range) score was 43.8 ± 18.3 (0-60) and 11 (65%) were classified as typically developing, 2 (12%) in the monitor zone, and 3 (18%) in the warning zone, with 1 parent not completing the ASQ (Figure 1 Panel A). Physiotherapists had an average of 8.5 ± 8.8 (1.0-30.0) years working with children with JIA. Most (54%; n=13/24) physiotherapists saw 10 or more children with JIA each month. When asking specifically about children with JIA aged 1-5 years, 46% (n=11) saw 2 to 5 children a month, followed by 25% (n=6) who saw 10 or more a month, then 17% (n=4) who saw 0-1, and finally 13% (n=3) who saw 6 to 9 a month.

**Figure 1.**
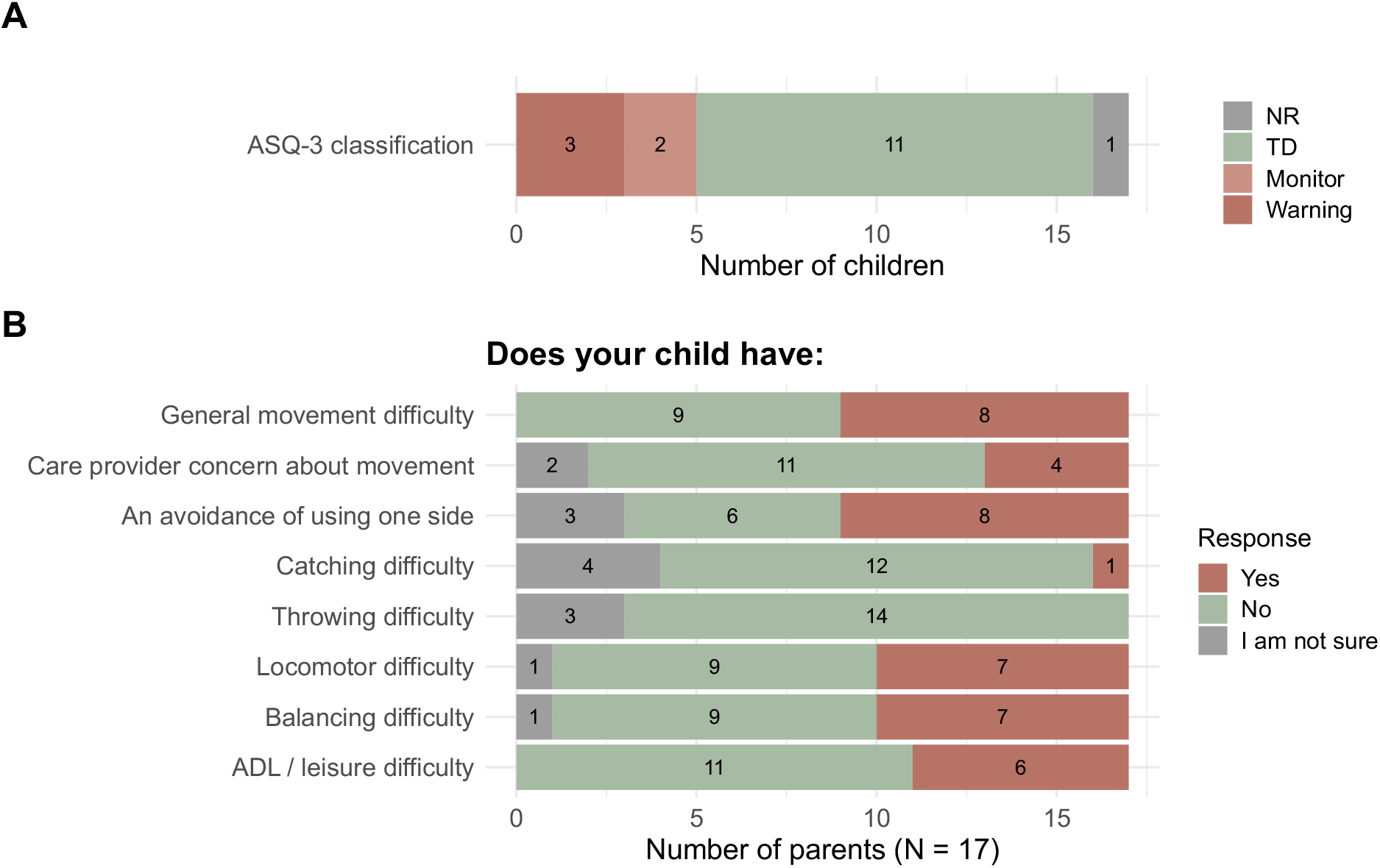
Child classification from the ASQ-3 (panel A) and parent responses from multiple choice questions in the questionnaire (Panel B). ASQ-3: ages and stages questionnaire – 3rd edition; NR: not reported; TD: typically developing. From left to right, Panel B responses are stacked as “I am not sure” (grey), “No” (green), and “Yes” (orange). ADL: activities of daily living.

### Quantitative analysis

From the perspective of parents, there was a nearly even split of parents who thought their children did have overall movements difficulties (47%; n=8) and those who did not (53%; n=9). Few reported that a care provider had expressed concern about their child’s movement skills (35%; n=6), but about half experienced their child avoiding the use of one side of their body (47%; n=8). Only 1 parent indicated that their child had difficulty with throwing or catching. Just under half of the parents (41%; n=7) indicated that their child had locomotor or balance difficulties. Finally, few (35%; n=6) parents felt that their children had difficulty with activities of daily living or leisure activities. Figure 1 panel B illustrates all parent responses. Supplementary material 2 shows parent responses separated by their child’s ASQ-3 classification (typically developing compared to monitor/warning zones). This shows that a higher proportion of parents with children in the monitor/warning zones report that their children have difficulty with all tasks, except catching and throwing, although there are only five children in the monitor/warning zone.
From the perspective of the physiotherapists, most (75%; n=18/24) described the movement skills of children with JIA they have worked with as worse than other children the same age, while the remaining 25% (n=6) described the movement skills as comparable to other children the same age. When asked to estimate the proportion of children with JIA who experience difficulty with movement skills, 42% (n=10) of physiotherapists reported that some (20-49%) children experience difficulty. Figure 2 panel A shows this detailed breakdown. When asked to estimate the proportion of children with JIA that avoid using one side or one part of the body, 42% (n=10) of physiotherapists reported that most (50-79%) children experience difficulty. Figure 2 panel B shows this detailed breakdown. Finally, Figure 3 provides a detailed breakdown of physiotherapist-reported frequency of children who have difficulty with upper body object control, lower body movement, and who experience functional limitations that impact their activities of daily living and/or leisure time activities.

**Figure 2.**
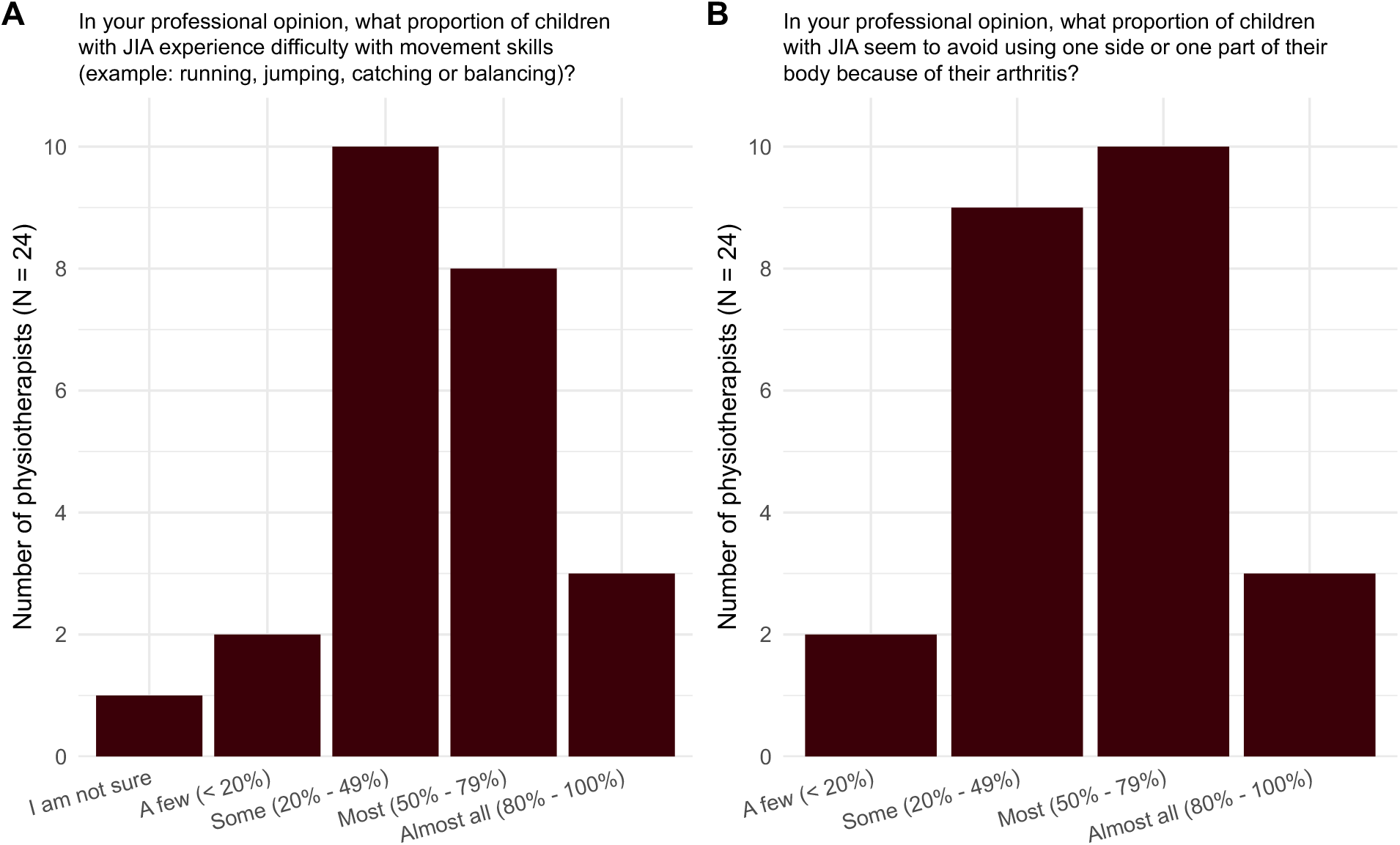
Physiotherapist estimates of proportion of children with JIA that experience difficulty with general movement skills (Panel A) and that avoid using one side (Panel B).

**Figure 3.**
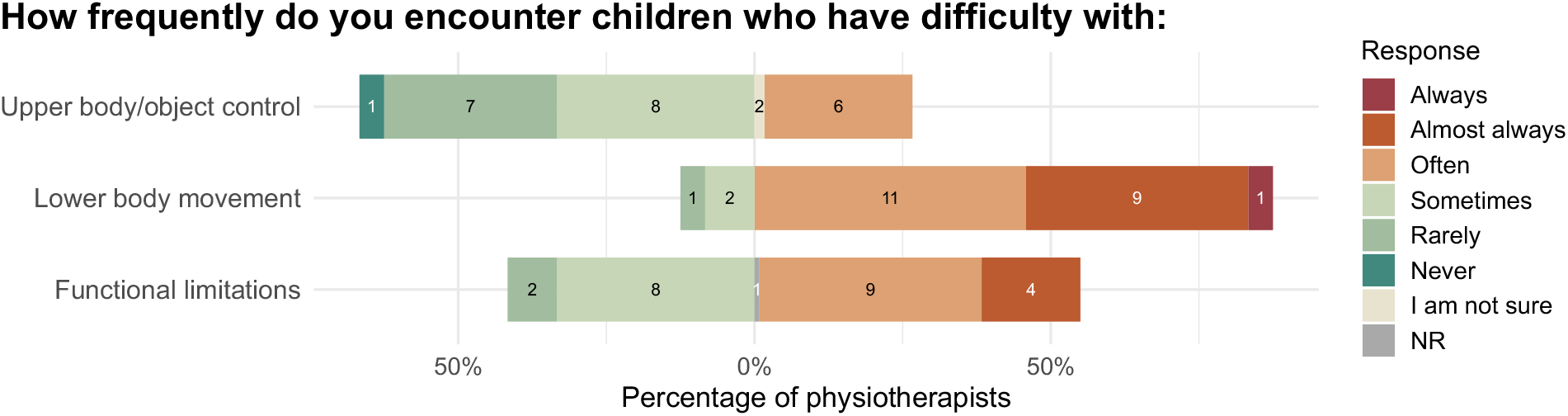
Physiotherapist estimates of frequency of children with JIA that experience difficulty with upper body movement, lower body movement, and functional limitations that impact their activities of daily living and/or leisure time activities. The x-axis shows the percentage of physiotherapists while numbers of physiotherapists per response are written on each bar segment.

### Qualitative analysis

In response to the open-ended text questions, 14/17 parents and 23/24 physiotherapists responded to at least 1 open-ended text prompt. Three general themes emerged regarding areas of concern from both parent and physiotherapists comments: (a) functional task difficulties; (b) clinical variability of JIA symptoms, treatments, and movement skills; and (c) psychosocial components of movement skill difficulties.

### Functional task difficulties

Participants often described situations in which a child with JIA requires additional support to complete a task or when a child changes their behaviour to perform a particular task. These tasks can include play, moving through space, as well as activities of daily living. One parent described that their child will, “avoid doing those activities that hurt and [call] for parents/teachers or help in those situations” (Participant 25). Similarly, a physiotherapist described that, “they will move in ways that protect their affected joints. There are lots of compensatory movement patterns.” (Participant 11). Participants describe how children may avoid using an affected limb and how this unilateral avoidance/preference is often maintained even after acute pain and inflammation are resolved. From the perspective of a parent, for example, “She also has developed a sort of skipping to avoid putting weight on her right ankle” (Participant 34). The adapted movement skills may become their “default” way of moving. Within the topic of functional tasks difficulties, participants identified seven specific tasks or areas with difficulty: (i) avoidance of using an affected limb, (ii) asymmetry, (iii) balance difficulties, (iv) limited endurance, (v) difficulty with transitions (e.g., sit to stand), (vi) hand/object control struggles for activities of daily living (e.g., gripping), and (vii) reduced strength/power.

### Clinical variability of JIA symptoms, treatments, and movement skills

Many participants described the variability of symptoms, treatments, and movement skills as children with JIA age. This variability can be in part due to flares, medication onset or changes, typical child movement development variability, as well as other co-occurring conditions/symptoms (e.g., muscle weakness, obesity). Physiotherapists noted that movement skill difficulties are, “usually more present if there is stiffness associated with arthritis,” (Participant 8) and often worsen, “during their flare ups” (Participant 14). A parent described that, “since being on medication [she] has improved a lot and is catching up to her age level in motor skills.” (Participant 12).
Parents and physiotherapists also mentioned the importance of being physically active and experimenting with new activities to help re-train movement skills. A parent described that, “Since taking medication she […] is catching up, but still behind. She is now trying new things on her own and realizing where her limbs are in space and what she can do. She will experiment with her movements.” (Participant 12). Similarly, from a physiotherapist, “Over time, I find most patients will “catch up” if they fall behind in skills - if they stay physically active in a variety of areas.” (Participant 3).

### Psychosocial components of movement skill difficulties

Participants described both parent and child perceptions and emotions towards movement skill difficulties. Parent emotions mentioned include distress at movement skill regression (e.g., child learned to walk, but then stopped during a flare) and concern around participation in physical activities inducing a flare. Child emotions mentioned include frustration in not being able to “keep up” with peers and fear in engaging in certain activities. From the perspective of a parent, “related to jumping, [he] does have difficulty. He tries with assistance [to] jump from a higher place, but he doesn’t feel comfortable doing it himself.” (Participant 36).
From the perspective of a physiotherapist, movement skill difficulties, “can persist in particular if parental anxiety or child develops preference for quiet activities and avoids sports or active play participation; ie if harder than their peers, may give up or parents may be anxious re: exacerbating arthritis and also facilitate lack of exposure to some play needed to develop skills with jumping etc.” (Participant 23).

## Discussion

This study aimed to understand the perceptions of physiotherapists and parents towards the movement skills of children with JIA. The quantitative analysis showed that about half of parents perceived their children to have movement skill difficulties and 75% of physiotherapists described the movement skills of children with JIA as worse than other children their age. Both parents and physiotherapists reported more difficulty with lower body movements (including locomotion) than upper body difficulties. From the qualitative analysis, participants identified three general themes: functional task difficulties; the clinical variability of JIA symptoms, treatments, and movement skills; and psychosocial components of movement skill difficulties.

Across both quantitative and qualitative responses, reference to lower limb difficulties were much more prevalent than upper limb. This is not entirely surprising as lower limb joints are more commonly impacted in JIA in this age group.^30^ In general, physiotherapists reported that a higher proportion of children experienced movement skill difficulties than parents, although this is almost certainly influenced by the fact that children who present with movement difficulties are more likely to be referred to physiotherapy. Only one parent indicated that their child had upper limb difficulties and physiotherapists reported that they observe upper limb difficulties sometimes (33% of physiotherapists) or often (25% of physiotherapists), with 30% reporting rarely. Despite this lower reporting of upper limb difficulties, future research into these impacts are warranted as upper limb involvement may result in higher levels of reported disability and worsened quality of life.^31^
From the perspective of both parents and physiotherapists, young children excel at compensating for painful movements and adapting how they complete activities to accomplish the desired task. Many also described an accelerated catch-up period once symptoms are controlled. However, participants further identified that children may struggle to re-learn skills even once the symptoms of pain and stiffness are improved or resolved. This reinforces the importance of referrals to physiotherapy and occupational therapy to help children gain strength, adopt appropriate movement patterns, and gain confidence in completing tasks, especially as they navigate the variability of their symptoms. Another area of future work to support children though the variability of symptoms could investigate supports provided to parents, such as a home-based movement skill toolkit. From the findings of this paper, such a toolkit should: (i) include both upper and lower body skill development, with more of an emphasis on lower body movement skills; (ii) provide educational information to parents around specific movement skill challenges that children with JIA may face and clear guidelines for when an activity should be stopped (addressing psychosocial concerns); (iii) provide support and resources specific to the variability of clinical symptoms (e.g., modifications of movement skills during and after a flare in a specific joint, addressing asymmetry); and (iv) play-based activities that focus on functional skills (e.g., specific skills integrated into an obstacle course).

While these findings provide unique insight into parent and physiotherapist perspectives towards the movement skills of children with JIA, this study has its limitations. First, the sample size is relatively small and we did not collect detailed demographic information such as ethnicity or socioeconomic status, which may limit the ability to generalize the findings to a wider population. Future research should be conducted with more participants to confirm these findings. Second, while using quantitative and qualitative methods of analysis helped strengthen our findings, the open-ended text responses elicited relatively short written responses, which may limit the depth of qualitative data. Future studies could employ interviews or focus groups to get a deeper understanding of parent and physiotherapists perceptions, as well as seeking the perspectives of other care providers and interest-holders. Finally, parents had mostly female (88%) children which, although aligning with the increased prevalence of JIA in females,^32^ may bias findings given the sex-based differences in movement skill development and parental perceptions of development and delays.^33^

## Conclusions

Movement skill development may be impacted when symptoms of JIA appear in the early years. Specifically, joint pain and stiffness may alter how children learn specific tasks, such as walking or drawing. About half of parents in this study perceived their children to have overall movement difficulties and 75% of physiotherapists described the movement skills of children with JIA as worse than other children their age. Participants identified three general themes including functional task difficulties, the clinical variability of JIA symptoms, treatments, and movement skills, and finally, psychosocial components of movement skill difficulties. Future work should continue to explore other avenues to support children’s movement skill development, including additional supports for parents, physiotherapists, and other care providers to monitor, flag, and intervene to promote movement skill development in the early years.

## Data Availability

The datasets analysed during the current study are available from the corresponding author on reasonable request.

## References

1. Palman J, Shoop-Worrall S, Hyrich K, McDonagh JE. Update on the epidemiology, risk factors and disease outcomes of Juvenile idiopathic arthritis. Best Practice & Research Clinical Rheumatology. 2018;32(2):206–222. doi:10.1016/j.berh.2018.10.004

2. Petty RE, Southwood TR, Manners P, et al. International League of Associations for Rheumatology classification of juvenile idiopathic arthritis: second revision, Edmonton, 2001. The Journal of Rheumatology. 2004;31(2):390–392.

3. Public Health Agency of Canada. Juvenile idiopathic arthritis in Canada. September 29, 2020. Accessed January 13, 2026. https://www.canada.ca/en/public-health/services/publications/diseases-conditions/juvenile-idiopathic-arthritis.html

4. Tuomi AK, Rebane K, Arnstad E, et al. Age at diagnosis as a prognostic factor in selected categories of juvenile idiopathic arthritis. RMD Open. 2025;11(2). doi:10.1136/rmdopen-2024-005369

5. van der Net J, van der Torre P, Engelbert RHH, et al. Motor performance and functional ability in preschool- and early school-aged children with Juvenile Idiopathic Arthritis: A cross-sectional study. Pediatric Rheumatology. 2008;6:2. doi:10.1186/1546-0096-6-2

6. Cools W, Martelaer KD, Samaey C, Andries C. Movement Skill Assessment of Typically Developing Preschool Children: A Review of Seven Movement Skill Assessment Tools. J Sports Sci Med. 2009;8(2):154–168.

7. Giddens JF. Concepts for Nursing Practice E-Book. Elsevier Health Sciences; 2023.

8. Gallahue DL, Ozmun JC, Goodway JD. Understanding Motor Development : Infants, Children, Adolescents, Adults. McGraw-Hill Education; 2012.

9. Gerber RJ, Wilks T, Erdie-Lalena C. Developmental Milestones: Motor Development. Pediatr Rev. 2010;31(7):267–277. doi:10.1542/pir.31-7-267

10. Yenil S, Gur Kabul E, Basakci Calik B, Kilbas G, Yuksel S. Investigation of motor skill in patients with juvenile idiopathic arthritis: A cross sectional study. Revista Colombiana de Reumatologia. 2025;32(1):36EP–42. doi:10.1016/j.rcreu.2023.11.005

11. Letts E, Savin K, Barbera L, et al. Motor skills of children with juvenile idiopathic arthritis: a scoping review. Zenodo. Preprint posted online May 20, 2026. doi:10.5281/zenodo.20314715

12. Bedard C, King-Dowling S, Timmons B, Ferro M. A Matched-Pair Analysis of Gross Motor Skills of 3-to 5-Year-Old Children With and Without a Chronic Physical Illness. Pediatric Exercise Science. 2025;37(1):75–80. doi:10.1123/pes.2023-0069

13. Morrison CD, Bundy AC, Fisher AG. The Contribution of Motor Skills and Playfulness to the Play Performance of Preschoolers. American Journal of Occupational Therapy. 1991;45(8):687–694.

14. Brandelli YN, Stone M, Westheuser V, et al. Parent Risk Perceptions, Physical Literacy, and Fundamental Movement Skills in Children With Juvenile Idiopathic Arthritis. Pediatric Physical Therapy. 2022;34(4):536–544. doi:10.1097/PEP.0000000000000948

15. Yuwen W, Lewis FM, Walker AJ, Ward TM. Struggling in the Dark to Help My Child: Parents’ Experience in Caring for a Young Child with Juvenile Idiopathic Arthritis. Journal of Pediatric Nursing: Nursing Care of Children and Families. 2017;37:e23–e29. doi:10.1016/j.pedn.2017.07.007

16. Di Ludovico A, La Bella S, Di Donato G, Felt J, Chiarelli F, Breda L. The benefits of physical therapy in juvenile idiopathic arthritis. Rheumatol Int. 2023;43(9):1563–1572. doi:10.1007/s00296-023-05380-9

17. Arbaciauskaite A, Dudoniene V. The Effect of Physiotherapy in the Treatment of Juvenile Idiopathic Arthritis. Reabilitacijos mokslai: slauga, kineziterapija, ergoterapija. 2016;1(14). doi:10.33607/rmske.v1i14.696

18. Monek B, Vitár D, Chrobot R, Orbán I, Kiss E, Poór G. AB1427-HPR The role of parents’ awareness in physical activity in children with juvenile idiopathic arthritis. Annals of the Rheumatic Diseases. 2018;77:1847. doi:10.1136/annrheumdis-2018-eular.5362

19. Stavropoulou M, Alexandra HP, Apostolou T, Iakovidis P, G Maria K, Manolis T. Parents’ opinion on pediatric physiotherapy and the physical therapy. Physiother Res Rep. 2020;3(1). doi:10.15761/PRR.1000129

20. Schroder N, Crabtree MJ, Lyall-Watson S. The Effectiveness of Splinting as Perceived by the Parents of Children with Juvenile Idiopathic Arthritis. British Journal of Occupational Therapy. 2002;65(2):75–80. doi:10.1177/030802260206500205

21. Rouster-Stevens K, Nageswaran S, Arcury TA, Kemper KJ. How do parents of children with juvenile idiopathic arthritis (JIA) perceive their therapies? BMC Complement Altern Med. 2008;8(1):25. doi:10.1186/1472-6882-8-25

22. Harris PA, Taylor R, Thielke R, Payne J, Gonzalez N, Conde JG. Research electronic data capture (REDCap)—A metadata-driven methodology and workflow process for providing translational research informatics support. Journal of Biomedical Informatics. 2009;42(2):377–381. doi:10.1016/j.jbi.2008.08.010

23. Juvenile Idiopathic Arthritis - Support and Resources - Cassie + Friends. Cassie and Friends Society. Accessed April 28, 2026. https://cassieandfriends.ca/

24. Letts E, King-Dowling S, Calotti R, Di Cristofaro N, Obeid J. Investigating the validity of the Ages and Stages Questionnaire to detect gross motor delays in a community sample of toddlers: A cross-sectional study. Early Human Development. 2023;187:105882. doi:10.1016/j.earlhumdev.2023.105882

25. Squires J, Twombly E, Bricker D, Potter L. ASQ-3 User’s Guide. Paul H. Brookes Publishing Co. Inc; 2009.

26. Çelen Yoldaş T, Özdel S, Karakaya J, Bülbül M. Developmental and Behavioral Problems of Preschool-Age Children with Chronic Rheumatic Diseases. J Dev Behav Pediatr. 2022;43(3):e162–e169. doi:10.1097/DBP.0000000000001007

27. Hsieh HF, Shannon SE. Three approaches to qualitative content analysis. Qualitative Health Research. 2005;15(9):1277–1288. doi:10.1177/1049732305276687

28. Dedoose. www.dedoose.com

29. Mayan MJ. Essentials of Qualitative Inquiry. 1st ed. Left Coast Press; 2009. https://www.routledge.com/Essentials-of-Qualitative-Inquiry/Mayan/p/book/9781598741070

30. Hemke R, Nusman CM, van der Heijde DMFM, et al. Frequency of joint involvement in juvenile idiopathic arthritis during a 5-year follow-up of newly diagnosed patients: implications for MR imaging as outcome measure. Rheumatol Int. 2015;35(2):351–357. doi:10.1007/s00296-014-3108-x

31. Buran S, Karaca NB, Tüfekçi MO, et al. Comparison of Disease Activity, Inflammatory Biomarker, Functionality, Participation and Biopsychosocial Status of Individuals with Jia According to the Presence of Upper Extremity Involvement. Annals of the Rheumatic Diseases. 2023;82(Suppl 1):2086–2087. doi:10.1136/annrheumdis-2023-eular.5130

32. Thierry S, Fautrel B, Lemelle I, Guillemin F. Prevalence and incidence of juvenile idiopathic arthritis: A systematic review. Joint Bone Spine. 2014;81(2):112–117. doi:10.1016/j.jbspin.2013.09.003

33. Liong GHE, Ridgers ND, Barnett LM. Associations between Skill Perceptions and Young Children’s Actual Fundamental Movement Skills. Percept Mot Skills. 2015;120(2):591–603. doi:10.2466/10.25.PMS.120v18 × 2

