## supplementary material 1 for "Parent and physiotherapist perceptions about movement skills of young children with juvenile idiopathic arthritis"

### Supplementary Material 1. Parent and physiotherapist surveys.

### Parent Survey

| | |
|---|---|
| Please identify your relationship to the child | ◯ Mother<br>◯ Father<br>◯ Other ______ |
| Please enter your child's date of birth | ______________________________<br>(DD-MM-YYYY) |
| What is your child's sex? | ◯ Male<br>◯ Female<br>◯ Other ______<br>◯ Prefer not to answer |
| Was your child born more than 3 weeks prematurely? | ◯ Yes<br>◯ No |
| How many weeks early was your child born? | ______________________________ |
| Please enter your child's year and month of diagnosis. | ______________________________<br>(If you can't remember the exact month, please enter the year.) |
| Which of your child's joints/body parts are affected by JIA? | _____________________________________<br>(example: left knee, right shoulder) |
| Has your child been diagnosed with any other medical conditions or developmental disorders? | ◯ Yes, please specify: ______ and year of diagnosis: ______<br>◯ No |
| What is your child's dominant hand/arm? | ◯ Left<br>◯ Right<br>◯ Both<br>◯ Unsure/Undetermined<br>(example: arm with which they hold a pencil) |
| What is your child's dominant leg? | ◯ Left<br>◯ Right<br>◯ Both<br>◯ Unsure/Undetermined<br>(example: leg your child kicks a ball with) |

### Fundamental Movement Skills

| | Yes | No | I am not sure |
|---|---|---|---|
| In your opinion, does your child have difficulty with movement skills (example: walking, running, jumping, catching or balancing)? | ○ | ○ | ○ |
| Has your child's care provider ever expressed concern about the development of your child's movement skills (example: walking, running, jumping, catching or balancing)? | ○ | ○ | ○ |
| In your opinion, does your child seem to avoid using a side or part of their body because of their arthritis (example: avoids using right wrist to protect it from pain)? | ○ | ○ | ○ |

### Upper Body Movement Skills

| | |
|---|---|
| In general, compared to children of the same age, does your child appear to avoid or have difficulty catching (with their left hand, right hand or both)? | ○ Yes<br>○ No<br>○ I am not sure |
| Do you think this is because of your child's arthritis? | ○ Yes<br>○ No |
| Please describe how arthritis has influenced your child's ability to catch | ________________________________ |
| In general, does your child appear to avoid or have difficulty throwing? | ○ Yes<br>○ No<br>○ I am not sure |
| Do you think this is because of your child's arthritis? | ○ Yes<br>○ No |
| Please describe how arthritis has influenced your child's ability to throw | ________________________________ |

### Lower Body Movement Skills

| | |
|---|---|
| In general, does your child appear to avoid or have difficulty doing locomotor activities involving their lower body such as walking, jumping or running? | ◯ Yes<br>◯ No<br>◯ I am not sure |
| Do you think this is because of your child's arthritis? | ◯ Yes<br>◯ No |
| Please describe how arthritis has influenced your child's ability to run, jump, walk | ______________________________ |
| In general, compared to children of the same age, does your child appear to avoid or have difficulty doing balancing activities using their lower body like balancing on one leg or kicking a ball? | ◯ Yes<br>◯ No<br>◯ I am not sure |
| Do you think this is because of your child's arthritis? | ◯ Yes<br>◯ No |
| Please describe how arthritis has influenced your child's ability to kick or balance | ______________________________ |

### Impact on Daily Living

| | |
|---|---|
| In general, compared to children of the same age, does your child appear to avoid or have difficulty doing activities of daily living and/or leisure-time activities like climbing the stairs or playing on the playground? | ◯ Yes<br>◯ No<br>◯ I am not sure |
| Do you think this is because of your child's arthritis? | ◯ Yes<br>◯ No |
| Please describe how arthritis has influenced your child's ability to engage in activities of daily living | ______________________________ |
| Is there anything else you'd like to share with us about your child's movement skills? | ______________________________ |

### Physiotherapist survey

How many years have you worked with children with juvenile idiopathic arthritis?

____________________
(# of years)

On average, how many children (of any age) with juvenile idiopathic arthritis do you care for in a month?

◯ 0 - 1
◯ 2 - 5
◯ 6 - 9
◯ 10 or more

How many children with juvenile idiopathic arthritis between the ages of 1 - 5 years do you care for in a month?

◯ 0 - 1
◯ 2 - 5
◯ 6 - 9
◯ 10 or more

#### Fundamental Movement Skills

On average, how would you describe the fundamental movement skills in most children you've treated with juvenile idiopathic arthritis?

◯ Better than other children their age
◯ Comparable to other children their age
◯ Worse than other children their age
◯ I am not sure

Please describe and provide specific examples, if possible

____________________

In your professional opinion, what proportion of children with JIA experience difficulty with movement skills (example: running, jumping, catching or balancing)?

◯ Almost all (80% - 100%)
◯ Most (50% - 79%)
◯ Some (20% - 49%)
◯ A few (< 20%)
◯ None
◯ I am not sure

In your professional opinion, what proportion of children with JIA seem to avoid using one side or one part of their body because of their arthritis?

◯ Almost all (80% - 100%)
◯ Most (50% - 79%)
◯ Some (20% - 49%)
◯ A few (< 20%)
◯ None
◯ I am not sure

#### Upper Body Movement Skills

Among the children with JIA that you work with, how frequently do you encounter children who have difficulty with upper body movement and object control (example throwing, catching)?

◯ Always
◯ Almost always
◯ Often
◯ Sometimes
◯ Rarely
◯ Never
◯ I am not sure

Please describe and provide specific examples

____________________

### Lower Body Movement Skills

| | |
|---|---|
| How frequently do you encounter children with JIA who have difficulty with lower body movement (example walking, running, kicking a ball, jumping/hopping) ? | ◯ Always<br>◯ Almost always<br>◯ Often<br>◯ Sometimes<br>◯ Rarely<br>◯ Never<br>◯ I am not sure |
| Please describe and provide specific examples | ________________________________ |

### Impact on Daily Living

| | |
|---|---|
| How frequently do you encounter children with JIA with functional limitations that impact their activities of daily living and/or leisure-time activities (example crawling, walking up the stairs, balancing, standing up from the floor, navigating obstacles in environment)? | ◯ Always<br>◯ Almost always<br>◯ Often<br>◯ Sometimes<br>◯ Rarely<br>◯ Never<br>◯ I am not sure |
| Please describe and provide specific examples | ________________________________ |
| Is there anything else you'd like to share with us about the movement skills of your young patients with JIA? | ________________________________ |
