## Supplementary material 2 for "Parent and physiotherapist perceptions about movement skills of young children with juvenile idiopathic arthritis"

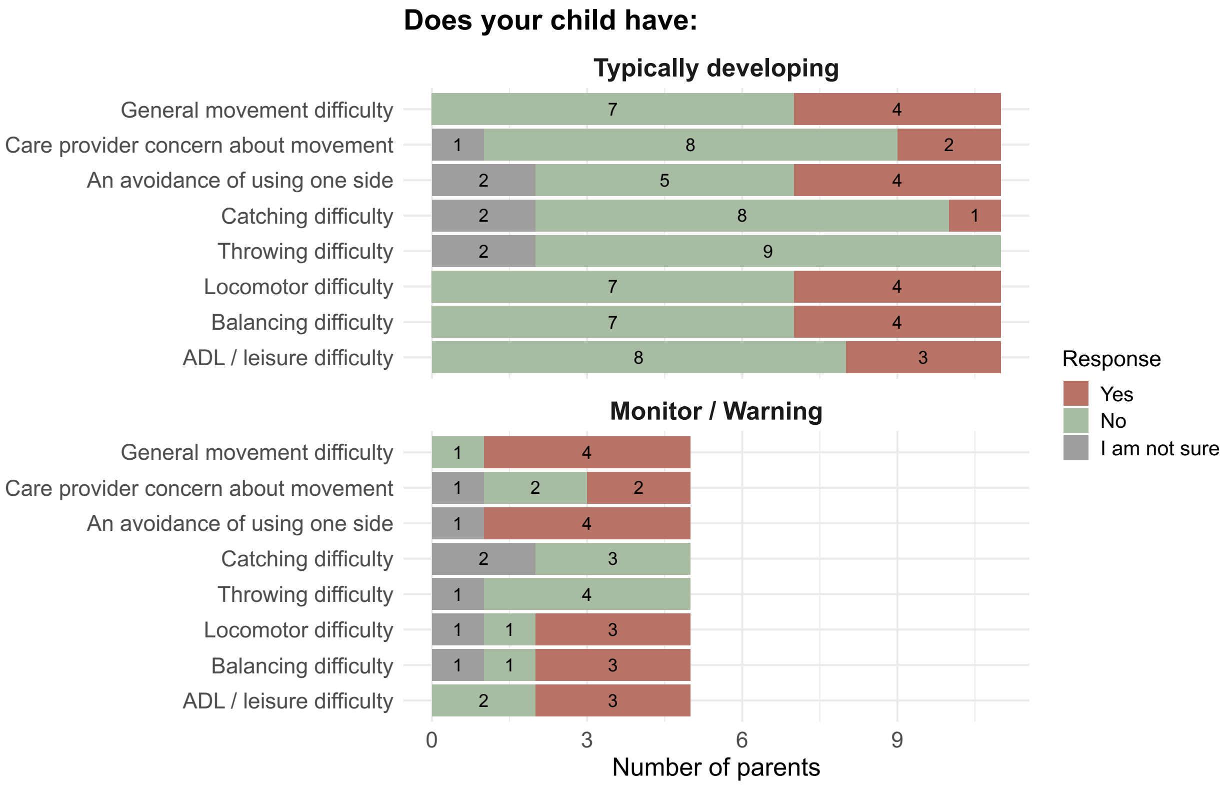


Parent responses from multiple choice questions in the questionnaire divided by their child's ASQ-3 classification. From left to right, responses are stacked as "I am not sure" (grey), "No" (green), and "Yes" (orange). ADL: activities of daily living; ASQ-3: ages and stages questionnaire – 3rd edition; NR: not reported; TD: typically developing.
